# Genetic and multi-omic prioritization implicates the CD33–SIGLEC locus in post-traumatic stress disorder

**DOI:** 10.64898/2026.09.23.26363743

**Authors:** Henghui Liu, Shuyan Xie, Zitao Chen, Qishan Wang

## Abstract

Genetic studies of post-traumatic stress disorder (PTSD) implicate immune regulation, yet linking peripheral immune-related proteins to brain molecular regulation and experimentally tractable targets remains challenging. Cellular and spatial genetic analyses were used to characterize the cellular and spatial genetic context of these traits. We then conducted bidirectional Mendelian randomization (MR) across 30 predefined Ig-like plasma proteins and 16 behavioral and neuropsychiatric traits. The forward MR screen identified four associations meeting the prespecified screening criteria, all of which underwent the same post-screening robustness assessment. CD33–PTSD showed the most consistent support across instrument definitions and applicable robust estimators and was therefore advanced as the prioritized candidate to regional genetic, brain molecular QTL, and functional analyses. Standard CD33 pQTL–PTSD colocalization did not support a simple shared single-variant model, while multi-signal analyses remained scientifically unresolved under the available LD reference. Meanwhile, plasma CD33 pQTL and cortical CD33 eQTL showed strong regional sharing (PP4=0.96468). Regional localization, cross-analysis candidate selection, and functional annotation further suggested a complex regulatory context across the CD33–SIGLEC region. Among 284 compounds with prior CD33 binding evidence that entered structure-assisted screening, 11 were retained as candidates for experimental follow-up. Overall, CD33–PTSD was the prioritized association identified by our screening; subsequent regional genetic, brain molecular QTL, and functional annotation analyses further delineated the complex regulatory context of the CD33– SIGLEC locus, while the CD33-centered structure-assisted exploration yielded 11 compound candidates for subsequent experimental validation.

## 1 Introduction

Post-traumatic stress disorder (PTSD) is shaped by genetic susceptibility, trauma exposure, and multisystem biological responses. A recent genome-wide association study (GWAS) meta-analysis of more than one million participants identified 95 genome-wide significant risk loci, with signals implicating stress responses, synaptic function, and endocrine and immune regulation; clinical and molecular studies also continue to link PTSD with peripheral inflammation and central neuroimmune states. Because peripheral and brain immune changes do not necessarily occur in parallel, GWAS loci or signals from a single tissue are insufficient to identify intervention-relevant effector molecules. Layered evaluation of genetic associations together with molecular phenotypes and cellular and tissue context is therefore needed. [1–2]

The plasma proteome provides a translational intermediate linking genetic variation, systemic molecular phenotypes, and potentially druggable proteins, and protein quantitative trait loci (pQTLs) can be used to systematically assess genetic associations between protein levels and complex traits. [3–4] MR alone, however, cannot establish that a protein directly mediates disease risk: linkage disequilibrium (LD), horizontal pleiotropy, differences between cis and trans instruments, and discordant regulation between plasma and disease-relevant tissues can all produce reproducible associations with non-simple mechanisms. Moving from pQTL screening to target interpretation therefore requires evaluation of instrument robustness, regional genetic sharing, and molecular QTL evidence from disease-relevant tissues.

Within this context, immunoglobulin-like receptors and related myeloid immune pathways have plausible neuroimmune relevance. CD33 (Siglec-3) is a myeloid cell-surface receptor of the CD33-related Siglec family and participates in regulating immune-cell activation thresholds, phagocytosis, and inflammatory regulation; in other neurological disorders, CD33 genetic variation and expression have been linked to monocyte and microglial function. This provides a biological rationale for systematically evaluating genetic associations between Ig-like plasma proteins, including CD33, and behavioral and neuropsychiatric traits, including PTSD, rather than prespecifying a particular protein–trait pair. [5–6]

We therefore used a hierarchical framework that moved from broad candidate screening to regional interpretation and translational exploration without prespecifying a single target protein. The study addressed three linked questions: first, which Ig-like plasma proteins are associated with PTSD and related behavioral and neuropsychiatric traits, as assessed by bidirectional MR and a symmetric robustness framework; second, whether prioritized associations receive support from regional genetic architecture and brain molecular regulation, while distinguishing simple shared signals from multi-signal or heterogeneous explanations; and third, whether the highest-priority protein candidate can be further resolved through functional annotation of candidate variants and translated into experimentally testable hypotheses by structure-assisted compound prioritization informed by existing binding evidence.

## 2 Materials and Methods

### 2.1 Study design and stepwise prioritization framework

We used a hierarchical, progressively convergent study design. From 184 neuro-related Olink assays reported by Repetto et al. [3], mapping and deduplication followed by InterPro IPR003599 (Ig_sub) [7] membership defined 30 candidate proteins, which were evaluated against 16 behavioral and neuropsychiatric traits. scDRS and gsMap were used to characterize the trait-level cellular and spatial genetic context.

Bidirectional MR was first performed across the full candidate space, after which associations meeting the prespecified screening criteria underwent the same post-screening robustness assessment. Downstream prioritization was based on consistency across instrument definitions and sensitivity analyses; from the regional-analysis stage onward, only CD33–PTSD was advanced. (Fig. 1)

**Figure 1.**
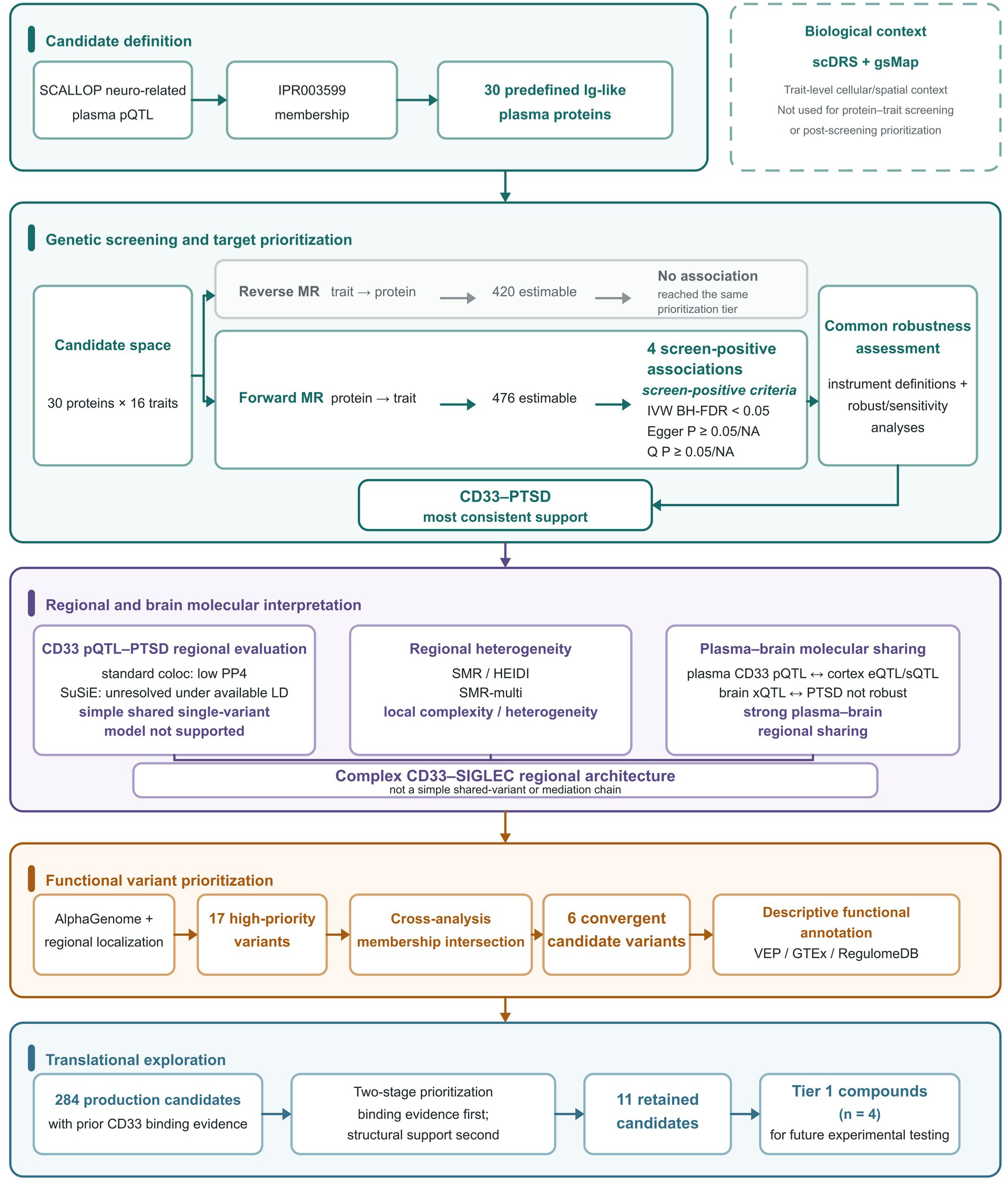
Study design and stepwise prioritization framework. Thirty predefined Ig-like plasma proteins, derived from the neuro-related Olink resource according to InterPro IPR003599 membership, were evaluated across 16 behavioral and neuropsychiatric traits; scDRS and gsMap provided trait-level cellular and spatial context only and were not used for protein–trait screening or post-screening prioritization. Bidirectional Mendelian randomization yielded 476 estimable forward associations and 420 estimable reverse associations. Four forward protein–trait combinations met the prespecified screening criteria, defined as IVW BH-FDR<0.05 together with MR-Egger intercept P≥0.05 or unavailable and Cochran’s Q P≥0.05 or unavailable; these associations then entered a common robustness assessment spanning different instrument definitions and robust or sensitivity analyses, and no reverse association reached the same prioritization tier. CD33–PTSD showed the most consistent support and was advanced to regional and brain molecular interpretation. Standard colocalization, SuSiE, SMR/HEIDI, SMR-multi, and plasma–brain molecular-sharing results were collectively more consistent with a complex CD33–SIGLEC regional genetic architecture than with a simple shared variant or mediation chain. AlphaGenome prioritization yielded 17 high-priority variants followed by regional localization; the cross-analysis membership intersection retained six convergent candidates, which were descriptively annotated using VEP, GTEx, and RegulomeDB. Translational exploration began with 284 production candidates with prior CD33 binding evidence and used a two-stage prioritization procedure: Stage 1 combined experimental binding potency, GNINA performance, and valid pocket entry to retain 28 candidates; Stage 2 further incorporated structural support, experimental potency, and PAINS-related rules to assign Tiers 1 –4, retaining 11 candidates and nominating four Tier 1 compounds for future experimental testing. NA, not available.

### 2.2 Candidate protein definition and genetic data sources

Candidate proteins were derived from the public resource of Repetto et al. (2024; DataShare DOI: 10.7488/ds/7522). [3] The 184 assays underwent name standardization, assay–gene mapping, and gene-symbol deduplication. Only assays that mapped uniquely and unambiguously to a gene were retained; ambiguous entries were not included in subsequent candidate definition. Thirty proteins were then fixed according to IPR003599 membership in the available DAVID/InterPro records. [7–8] The complete candidate definition and assay–gene mapping are provided in Supplementary Table S3.

Plasma pQTLs were used as protein exposures and GWAS data for 16 behavioral and neuropsychiatric traits as outcomes. [1,9–20] The datasets were predominantly of European ancestry, and regional analyses used GRCh37/hg19 coordinates. Phenotype definitions, sample sizes, effect/statistic scales, and data sources are provided in Supplementary Table S1.

### 2.3 Cellular and spatial genetic context of behavioral and neuropsychiatric traits

Cellular context was evaluated using MAGMA [21]/scDRS [22], whereas spatial context was assessed with gsMap [23] using tissue annotations from two MOSTA E16.5 sections [24], followed by cross-section aggregation [25]. BH false discovery rate (BH-FDR) [26] correction was applied jointly across retained trait–tissue gsMap combinations. Full methods, thresholds, and reproducibility boundaries are provided in Supplementary Methods S1.

### 2.4 Bidirectional MR screening and robustness-based prioritization

Formal bidirectional MR screening covered 30 candidate proteins and 16 behavioral and neuropsychiatric traits. Forward protein→trait analysis comprised 30×16=480 prespecified combinations, of which 476 were estimable; reverse trait→protein analysis covered 14 traits with usable inputs, yielding 420 estimable combinations. Forward analyses used pQTL instruments at P<1×10⁻⁵, whereas reverse analyses used GWAS variants at P<5×10⁻⁸. Single-SNP analyses used the Wald ratio; for multiple SNPs, inverse-variance weighted (IVW) MR [27] was the primary estimator, with MR-Egger [28], weighted median [29], and mode-based methods [30] also calculated. Instrument strength, Cochran Q heterogeneity, and the MR-Egger intercept were recorded.

BH-FDR correction [26] was applied to all formal MR results within each direction×method family. IVW BH-FDR<0.05 was the primary association-level significance threshold. Results meeting this threshold together with MR-Egger intercept P≥0.05 or unavailable and IVW Cochran-Q P≥0.05 or unavailable were classified internally as High and are reported here as associations meeting the prespecified screening criteria. BH-FDR<0.10 contributed to a supporting evidence tier; Moderate and Suggestive were supporting classifications defined by the full prespecified criteria in Supplementary Methods S2 and did not define associations meeting the prespecified screening criteria. The four associations meeting the prespecified screening criteria then underwent the same post-screening robustness framework, including cis-only, stringent-all, stringent-cis, leave-one-out, MR-PRESSO [31], MR-RAPS [32], and heterogeneity/horizontal-pleiotropy assessments. These analyses evaluated sensitivity to instrument definition and estimator choice. Downstream prioritization considered consistency across instrument definitions and sensitivity analyses. LD clumping, harmonization, method applicability, and classification thresholds are detailed in Supplementary Methods S2.

### 2.5 Regional genetic evaluation of the CD33–PTSD association

The formal standard colocalization analysis was restricted to plasma CD33 pQTL and PGC PTSD Freeze 3. coloc.abf [33] used CD33±500 kb as the primary window, with ±250 kb and ±1 Mb windows and alternative p12 priors as sensitivity analyses. This analysis was used to evaluate regional genetic sharing under a single-signal assumption. Because original beta/SE/OR values were unavailable for PGC PTSD, signed Z, allele frequency, and effective sample size (NEFF) were used to construct working-scale effects; this working scale was used only for regional comparison and was not reported or transformed as log-odds/OR.

Multi-signal analysis first used ordinary/reference SuSiE-RSS [34–35] and then sensitivity models accounting for the finite reference and summary–LD mismatch. Signal-level coloc.susie [36] results were considered scientifically interpretable only when the primary mismatch-aware models for both datasets in the same window passed the prespecified RELIABLE gate. PIP, credible-set, and PP.H4 outputs that did not pass this gate were retained only as diagnostic records and were neither interpreted as successful fine-mapping nor classified as a simple negative result. As complementary frameworks addressing different statistical questions, target-SMR/HEIDI [37] and SMR-multi [38] evaluated the regional relationship between CD33 cis-pQTL and PTSD on the PGC signed-Z/FREQ/NEFF working scale; SMR-multi was used as a set-level regional sensitivity analysis. Model settings, reliability gates, LD-processing procedures, and SMR parameters are provided in Supplementary Methods S3.

### 2.6 Regional sharing of peripheral and brain molecular regulation of CD33

Brain molecular QTL analyses separately evaluated regional sharing between plasma CD33 pQTL and BrainMeta v2 [39] cortical CD33 eQTL/nine sQTL molecular events, and between the corresponding cortical xQTLs and PTSD. Both analysis families consisted of independent pairwise colocalizations [33] and did not constitute a three-variable mediation model. Input data, regions, priors, and quality control are detailed in Supplementary Methods S4.

### 2.7 Functional prioritization, cross-analysis candidate selection, and locus annotation of CD33–PTSD candidate variants

AlphaGenome [40] was used for multimodal sequence-based functional prediction of CD33 –PTSD instrument variants after harmonizing REF/ALT orientation with the pQTL effect direction. Within the study, only variants with resolvable allele orientation and predicted direction concordant with the pQTL direction were ranked: the top 5% by absolute direction-adjusted predicted effect were classified as high priority, and the remaining variants within the top 15% were classified as moderate priority. This ranking was used to identify prioritized variants for subsequent focused functional annotation.

AlphaGenome-prioritized variants were then annotated independently for gene and regional position to describe their spatial relationship to CD33 and the neighboring SIGLEC region.

MR instrument SNPs, formal target-SMR records, effective SMR-multi members, and AlphaGenome high-priority SNPs were then intersected to narrow the candidate set for focused interpretation. This intersection was used to define the candidate set for subsequent in-depth annotation.

The fixed candidates were further annotated using VEP [41], GTEx v8 [42], and RegulomeDB v2 [43], together with targeted CD33 eQTL/sQTL queries. These analyses were used to describe the potential functional and regulatory context of the candidate variants. Locus definitions, membership of the six SNPs, coordinates, and query rules are provided in Supplementary Methods S5.

### 2.8 Experimental-binding-evidence-informed and structure-assisted prioritization of CD33 compounds

The CD33 small-molecule analysis was positioned as an independent post-prioritization translational exploration. Candidate ranking was anchored primarily by previously reported recombinant human CD33 competitive-binding EC50 values [44–45], with structural prediction [46] providing secondary support only. Docking scores, predicted contacts, and pocket occupancy were treated as computational descriptors rather than measured affinity or confirmed binding modes.

Known-drug and target-development context was curated separately by source: ChEMBL [47] and available licensed DrugBank records [48] were used for drug and compound background, whereas TTD [49], Thera-SAbDab [50], ClinicalTrials.gov, and related resources were used for target-development context; antibodies and antibody–drug conjugates were excluded from small-molecule docking. Formal small-molecule candidates came from the registered BindingDB/patent source set [44–45], comprising 322 standardized compound entities with unambiguous chemical structures and documented recombinant human CD33 competitive-binding activity. Of these, 284 achieved exact entity-level correspondence with the GNINA production library through standardized compound identifiers and entered structure-assisted ranking; the remaining 38 did not enter the production library because ligand preparation failed at ETKDGv3 [51] conformer generation.

Before formal GNINA production screening [46], native-pose recovery was evaluated in three cognate CD33 systems: 7AW6/FVP [52], 5J06/3′-sialyllactose [53], and 5J0B/6′-sialyllactose [54]. Redocking was used to assess native-pose recovery. Production screening used the 5J06 chain-B receptor and the binding pocket defined by 3′-sialyllactose. Because redocking performance was inconsistent across the three reference systems, GNINA was consistently treated only as secondary structural support layered onto the existing experimental binding evidence.

Candidates were evaluated using an explicit two-stage procedure without a weighted aggregate score. Stage 1 used experimental EC50, rank-1 GNINA scores [46], and valid pocket entry to assign the 284 production candidates to Groups A/B/C/D; 28 candidates in Groups A/B/C advanced to the next stage, whereas Group D was not advanced. Physicochemical properties and pharmacological context were considered only after Stage 1. Stage 2 combined structural support, PAINS [55], and related rules [56] to assign Tiers 1–4; Tiers 1–3 were retained and Tier 4 was not retained. The production docking box, GNINA parameters, grouping thresholds, structural-support criteria, tiering rules, and reproducibility limitations are detailed in Supplementary Methods S6.

### 2.9 Use of generative AI tools

Generative artificial intelligence tools, including OpenAI ChatGPT, were used to assist with the drafting and debugging of analysis and plotting code and with language editing of the manuscript. All AI-assisted code, text, and analytical suggestions were independently reviewed and verified by the authors before use. Generative AI tools were not used to autonomously determine the study design, data selection, parameter thresholds, quality-control criteria, candidate prioritization, or scientific conclusions; these decisions were made and finally reviewed by the authors. The authors take full responsibility for the accuracy, integrity, and conclusions of the work.

## 3 Results

### 3.1 Cellular and spatial genetic analyses indicate a neuroimmune context for behavioral traits

As context for subsequent genetic screening, scDRS and gsMap were used to characterize GWAS signals for the 16 behavioral and neuropsychiatric traits at cellular and spatial scales, collectively implicating immune, vascular/stromal, neural, and barrier-related contexts.

In scDRS, myeloid/lymphoid immune, vascular/stromal, and barrier-related cell types recurred across multiple traits and tissues. In gsMap, Brain, Spinal cord, and Meninges reached global BH-FDR<0.05 for 14 of 16 traits, whereas Choroid plexus, Dorsal root ganglion, Sympathetic nerve, and Inner ear reached this threshold for 13 of 16 traits. These results summarize the neuroimmune/barrier genetic context of the 16 traits; subsequent analyses proceeded to independent protein–trait genetic screening (Fig. 2a–d). Complete scDRS and gsMap results are provided in Supplementary Tables S4a and S4b, respectively.

**Figure 2.**
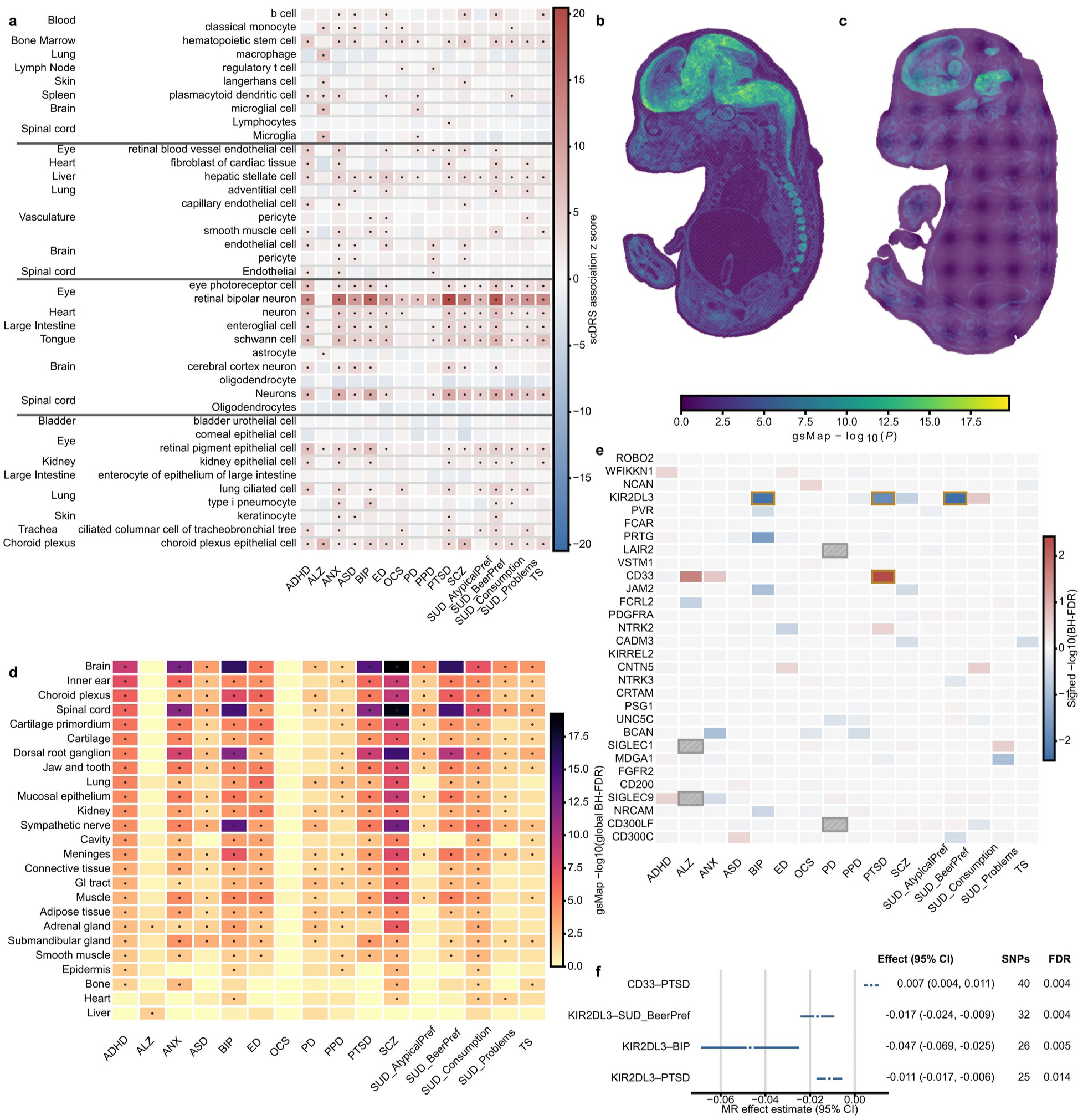
Cellular and spatial genetic context and forward MR screening of predefined Ig-like plasma proteins. a, scDRS association z scores for selected cell types across the 16 behavioral and neuropsychiatric traits; dots indicate cell type–trait combinations meeting the prespecified scDRS support criteria. Representative cell types are arranged from top to bottom by predefined broad cell categories: immune/hematopoietic, vascular/stromal, neural/glial, and barrier/epithelial; dark horizontal lines mark boundaries between cell categories. b,c, PTSD gsMap spatial-association maps in two MOSTA E16.5 embryonic sections; color indicates spatial −log10(P). d, gsMap trait–tissue summary across the 16 traits, shown as −log10(global BH-FDR); dots indicate trait–tissue combinations with global BH-FDR<0.05. e, Formal forward MR screening matrix for 30 predefined Ig-like plasma proteins across 16 traits. Color shows signed −log10(BH-FDR) according to IVW effect direction; hatched cells denote the four non-estimable combinations (LAIR2–PD, SIGLEC1–ALZ, SIGLEC9–ALZ, and CD300LF–PD), and outlined cells mark the four associations meeting the prespecified screening criteria. OCS (obsessive-compulsive symptoms) is used as the formal display label. f, IVW effect estimates and 95% confidence intervals for the four associations meeting the prespecified screening criteria, together with instrument-SNP counts and BH-FDR values. These four associations then entered the same post-screening robustness assessment. BH-FDR, Benjamini–Hochberg false discovery rate; IVW, inverse-variance weighted; MR, Mendelian randomization; PTSD, post-traumatic stress disorder.

### 3.2 Bidirectional MR and robustness analyses prioritize the CD33–PTSD association

The formal forward MR screen covered 30 predefined Ig-like plasma proteins and 16 behavioral and neuropsychiatric traits, yielding 30×16=480 prespecified protein–trait combinations, of which 476 had primary estimates; the four non-estimable combinations were LAIR2–PD, SIGLEC1–ALZ, SIGLEC9–ALZ, and CD300LF–PD. Reverse trait→protein analysis covered 14 traits with usable inputs and yielded 420 estimable combinations. The forward screen identified four associations meeting the prespecified screening criteria: CD33– PTSD, KIR2DL3–BIP, KIR2DL3–PTSD, and KIR2DL3–SUD_BeerPref. No reverse association reached the same prioritization tier. All estimable bidirectional MR results are provided in Supplementary Table S5; the complete 480-cell forward screening matrix, including the four non-estimable combinations, is retained in Source Data Fig2E_MR_screen. (Fig. 2e,f)

The four associations then underwent the same post-screening robustness assessment. Advancement to downstream analyses was based on a qualitative prioritization of consistency across this common robustness panel rather than on an additional prespecified statistical significance threshold. CD33–PTSD showed the most consistent support across instrument definitions and applicable robust estimators. The three KIR2DL3 associations depended more strongly on the full instrument sets containing both cis- and trans-pQTLs and showed weaker cis-only support; KIR2DL3–PTSD was particularly sensitive to a single locus. CD33–PTSD was therefore advanced to regional genetic, brain molecular QTL, and functional analyses. This post-screening prioritization is shown in Fig. 3a–c. Complete post-screening robustness results for the four associations are provided in Supplementary Tables S6a–S6h.

**Figure 3.**
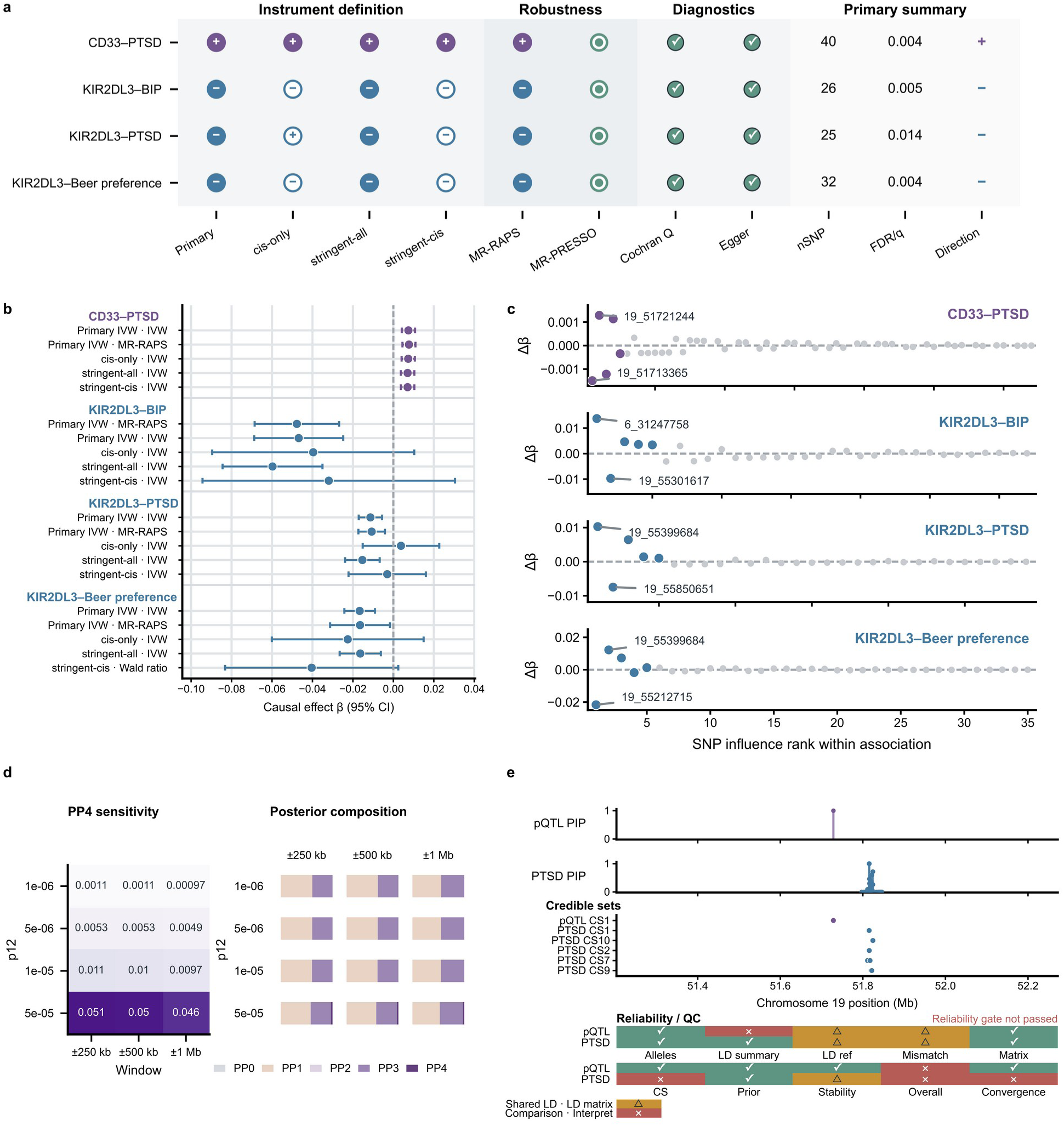
Common post-screening robustness assessment and regional evaluation of the four associations meeting the prespecified screening criteria. a, Summary of the primary, cis-only, stringent-all, and stringent-cis instrument definitions, applicable MR-RAPS and MR-PRESSO analyses, Cochran-Q and MR-Egger diagnostics, and the primary SNP count, FDR/q value, and effect direction for each association. Colors distinguish the different associations, while effect direction is indicated by “+”/“−”; filled/open symbols distinguish nominally significant from non-significant displayed MR estimates where applicable. In the MR-PRESSO column, entries denote outlier-analysis status rather than an effect estimate; when no outlier was identified, no outlier-corrected beta was generated. Unavailable or non-estimable analyses are shown with the corresponding status. b, Effect estimates and 95% confidence intervals for each association across primary IVW, primary MR-RAPS, and available alternative instrument definitions. c, Ranked SNP-influence display from leave-one-out analyses for the four associations; Δβ shows the change in the association estimate relative to the full primary estimate, with the most influential loci labeled. d, Standard plasma CD33 pQTL–PTSD colocalization across ±250-kb, ±500-kb, and ±1-Mb windows and alternative p12 values, showing PP4 sensitivity and the full PP0–PP4 posterior composition. Low PP4 did not support a simple shared single-variant model. e, Diagnostic SuSiE-RSS PIP and credible-set outputs with the corresponding reliability/QC summary. Under the available finite external LD reference and summary–LD matching conditions, the reliability gate was not passed; these outputs are diagnostic only and are not interpreted as successful fine-mapping, evidence of sharing, or a simple negative multi-signal result. FDR, false discovery rate; IVW, inverse-variance weighted; LD, linkage disequilibrium; PIP, posterior inclusion probability.

However, MR prioritization of CD33–PTSD was not supported by a simple shared genetic model. Standard pQTL–PTSD colocalization showed low PP4 across windows and priors. Although multi-signal SuSiE analyses were performed, the reliability gate was not passed under the available finite external LD reference and summary–LD matching conditions; because this gate was not passed, the corresponding PIP, credible-set, and coloc.susie outputs were not interpreted at the signal level. This discordance between MR and disease colocalization prompted further evaluation of regional complexity and molecular-QTL sharing across the CD33 –SIGLEC locus. (Fig. 3d,e) Complete standard colocalization and SuSiE diagnostic results are provided in Supplementary Tables S7a–S7j.

### 3.3 Regional genetic and brain molecular QTL analyses indicate a complex regulatory architecture at the CD33–SIGLEC locus

To further examine this discordance, target-SMR/HEIDI evaluated the local CD33–PTSD genetic architecture on the PGC signed-Z/FREQ/NEFF working scale, which is not the original log-odds or OR scale. The top cis-pQTL, rs2455069, reached nominal significance only (P=0.0380, BH-FDR=0.418), while HEIDI heterogeneity was significant (P=7.32×10⁻⁵), which did not support a simple single-signal shared model. SMR-multi yielded an exploratory set-level association (P=0.006622), suggesting that more complex local genetic structure should be considered, but did not imply SNP-level significance or multi-SNP causal effects. (Fig. 4d,e) Complete target-SMR/HEIDI and SMR-multi results are provided in Supplementary Tables S7k and S7l, respectively.

**Figure 4.**
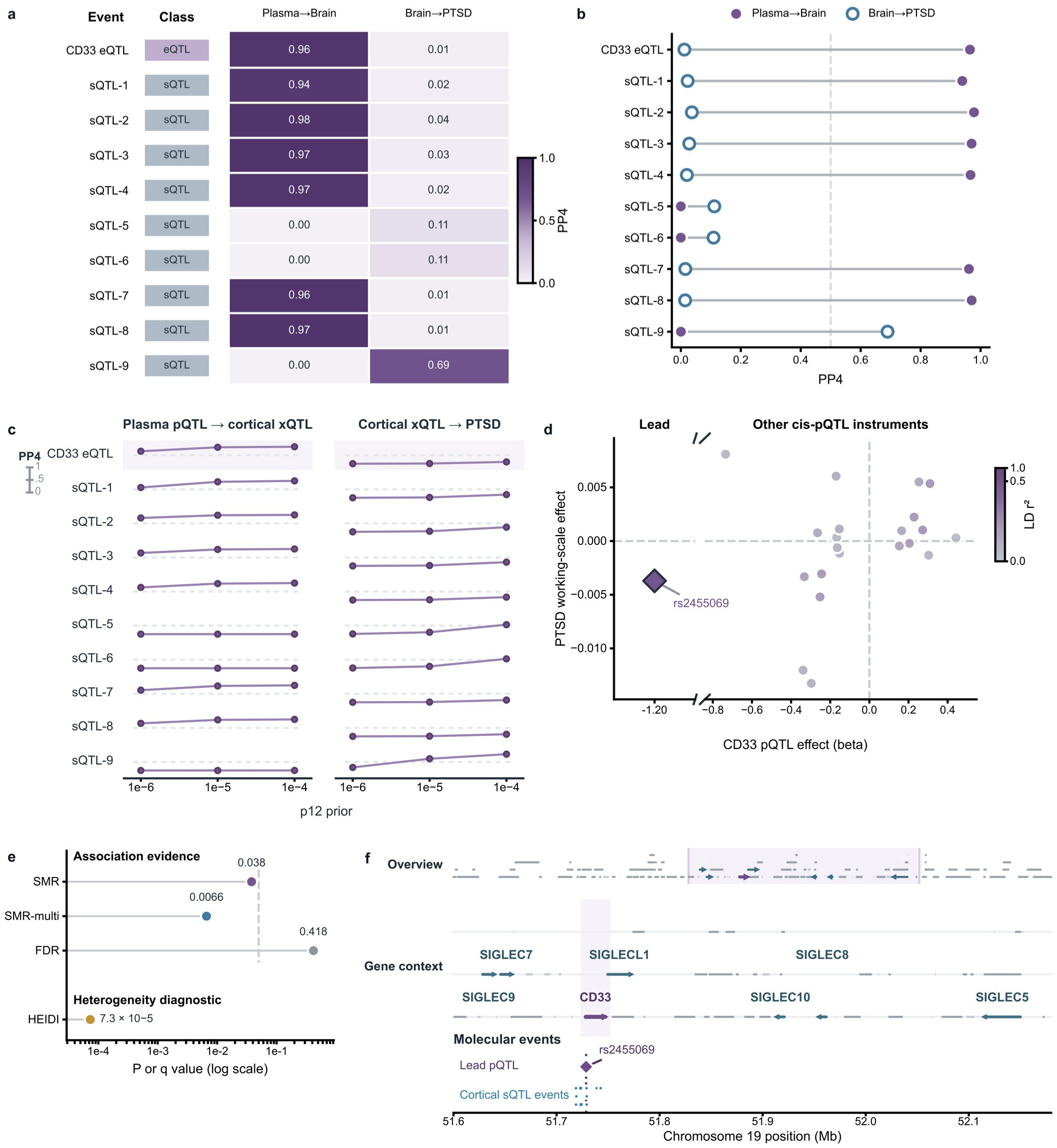
Brain molecular-QTL sharing and regional genetic architecture at the CD33–SIGLEC locus. a, Primary PP4 values for two separate families of pairwise colocalization: plasma CD33 pQTL with cortical CD33 eQTL and nine sQTL molecular events (Plasma→Brain), and the corresponding cortical xQTLs with PTSD (Brain→PTSD). b, Paired display of the same primary PP4 values, emphasizing the contrast between plasma–brain and brain–PTSD regional sharing. c, PP4 sensitivity across p12=10⁻⁶, 10⁻⁵, and 10⁻⁴ for both pairwise families. The nine sQTL records are molecular events rather than nine independent genetic signals, and the two pairwise families cannot be combined into a pQTL→brain xQTL→PTSD mediation or causal chain. d, CD33 cis-pQTL effect estimates versus PTSD working-scale effects, with the prespecified lead cis-pQTL rs2455069 highlighted and other cis-pQTL instruments colored by LD rZ to the lead. The PTSD axis is based on the signed-Z/FREQ/NEFF-derived working scale and is not interpreted as a log-odds effect. e, Target-SMR/HEIDI and exploratory SMR-multi summary. Target-SMR yielded P=0.038, BH-FDR=0.418, and HEIDI P=7.3×10⁻⁵; SMR-multi yielded a set-level P=0.0066. These results do not support a simple shared single-variant model and do not imply SNP-level significance or mediation. f, Regional overview of the lead pQTL, cortical molecular events, and gene context across the predefined CD33–SIGLEC region in GRCh37/hg19 coordinates. BH-FDR, Benjamini–Hochberg false discovery rate; eQTL, expression quantitative trait locus; HEIDI, heterogeneity in dependent instruments; LD, linkage disequilibrium; pQTL, protein quantitative trait locus; sQTL, splicing quantitative trait locus; SMR, summary-data-based Mendelian randomization; xQTL, molecular quantitative trait locus.

Cortical CD33 eQTL and sQTL signals did not show robust colocalization with PTSD. In contrast, plasma CD33 pQTL showed strong regional sharing with cortical CD33 eQTL (PP4=0.96468), and 6 of 9 sQTL molecular events also showed strong sharing (PP4>0.93). Thus, peripheral CD33 protein regulation shared regional genetic signals with cortical expression/splicing, but this sharing did not robustly extend to the PTSD disease signal. (Fig. 4a– c) Complete event-level results for the two families of pairwise colocalization are provided in Supplementary Tables S8a and S8b, respectively.

Taken together, the MR, disease colocalization, SMR/HEIDI, and brain molecular QTL findings were more consistent with a genetically complex CD33–SIGLEC region characterized by multiple signals and cross-tissue regulation than with a single shared CD33 variant model. We therefore next focused on functional prioritization, regional localization, and targeted annotation of candidate variants. (Fig. 4a–f)

### 3.4 Multimodal functional prediction and cross-analysis candidate annotation further resolve the CD33–SIGLEC region

Given the complexity of the local genetic architecture, AlphaGenome was used to assess potential regulatory effects of CD33–PTSD instrument variants. Predicted signals for high-priority variants arose mainly from CAGE, RNA-seq, and selected splicing and chromatin modalities. Among the 17 high-priority variants, one was located within the CD33 gene body, an additional 12 were within CD33±500 kb, and 13 were within the predefined CD33–SIGLEC region. Overall, high-priority variants identified by sequence-based functional prioritization were spatially concentrated primarily in the CD33–SIGLEC neighborhood and displayed multimodal regulatory features, consistent with the complex local structure suggested by the preceding regional genetic analyses and providing a functional basis for subsequent focused variant selection. (Fig. 5a–c)

**Figure 5.**
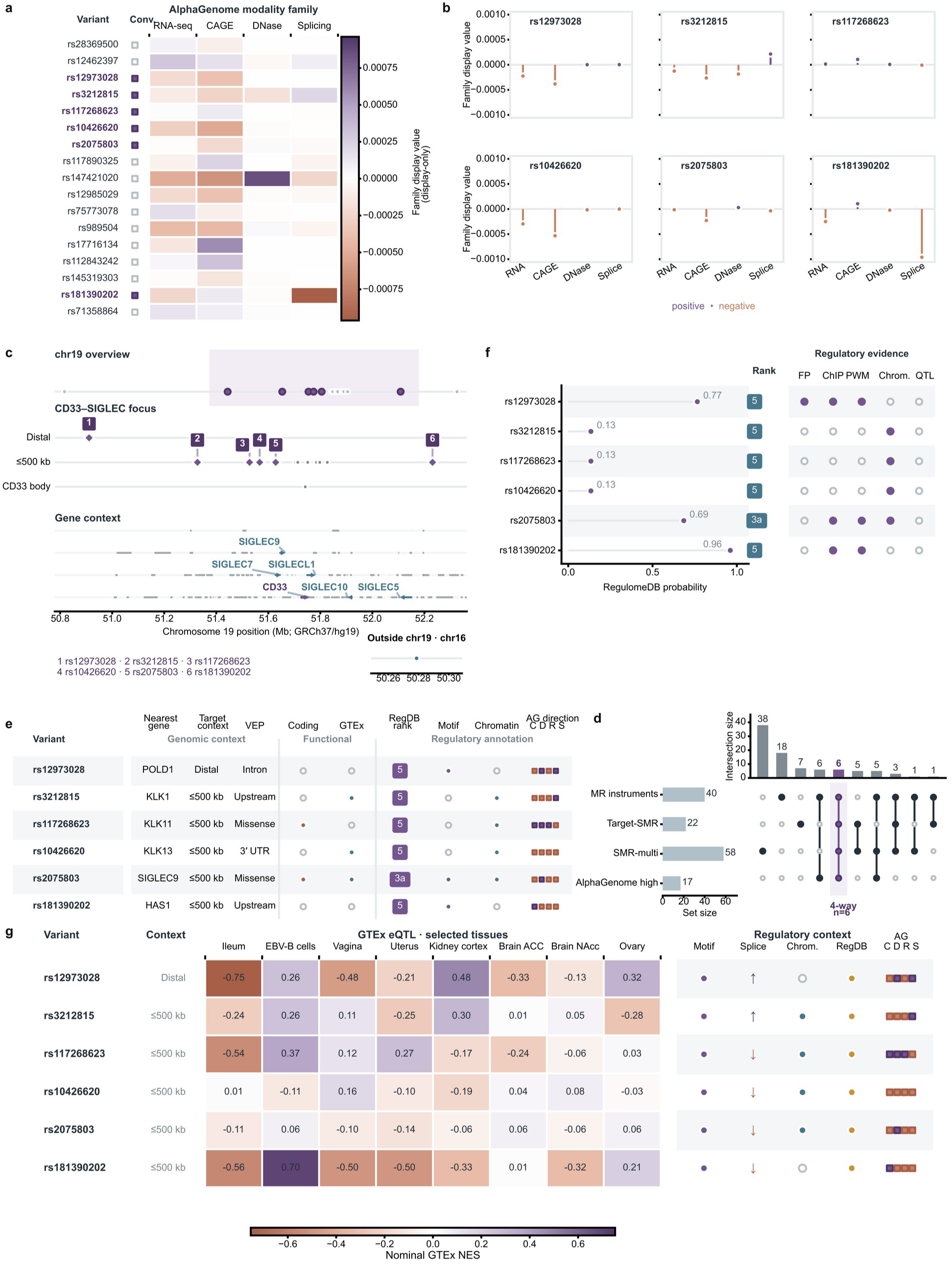
Multimodal functional prioritization and focused annotation of six convergent CD33– PTSD candidate variants. a, Direction-adjusted AlphaGenome family-level predicted effects for the 17 high-priority variants across RNA-seq, CAGE, DNase, and splicing modality families; the convergence marker identifies members of the six-variant intersection. b, Signed family-level AlphaGenome values for each of the six convergent candidate variants; purple and orange indicate positive and negative predicted effects, respectively. c, Genomic localization of the 17 high-priority variants and the six numbered candidates across chromosome 19, the predefined CD33–SIGLEC locus, CD33±500 kb, and the CD33 gene body, with regional gene context. d, UpSet-style membership display for MR instruments, target-SMR records, effective SMR-multi members, and the AlphaGenome high-priority set. The strict four-set intersection contains six convergent candidate variants; this represents membership overlap only and does not constitute a joint statistical test, a credible set, independent confirmation, or evidence of shared causal variants. e, Focused annotation matrix for the six candidates, including nearest-gene and CD33-region context, VEP consequence/coding status, source-defined GTEx annotation, RegulomeDB rank, regulatory-context indicators, and AlphaGenome direction across modality families. f, RegulomeDB probability, rank, and evidence-category indicators for the six candidates. g, GTEx v8 nominal eQTL NES for CD33 across selected tissues for the six candidate variants, together with regulatory-context indicators. The NES values shown derive from targeted six-variant×CD33 queries; all are nominal associations and did not meet source-defined significance criteria. All functional and regulatory annotations are descriptive and do not establish a unique causal gene. eQTL, expression quantitative trait locus; NES, normalized effect size; VEP, Variant Effect Predictor.

After AlphaGenome-based functional prioritization and regional localization, MR instruments, formal target-SMR records, effective SMR-multi members, and AlphaGenome high-priority members were intersected, yielding six convergent candidate variants, five of which were located within the CD33–SIGLEC region. This intersection was used to define the candidate set for subsequent focused functional annotation. (Fig. 5d)

The six candidates were then integrated with descriptive VEP, GTEx, and RegulomeDB regulatory annotations and targeted CD33 eQTL/sQTL queries. VEP/GTEx annotations implicated neighboring genes including POLD1, members of the KLK family, SIGLEC9, and HAS1. Targeted GTEx queries across the six variants×CD33 identified no direct CD33 eQTL/sQTL records defined as significant by the source resource; neither failure to meet source-defined significance nor isolated query failures were interpreted as excluding true CD33 regulation. rs2075803 also had a coding consequence in SIGLEC9. RegulomeDB annotations for the six variants were used only as descriptive regulatory context and were not treated as independent functional validation or as evidence establishing causality. Overall, annotation of the six variants did not converge on a single CD33 regulatory pathway; together with the regional genetic results, the overall evidence was compatible with regional architecture involving complex LD, multiple signals, multi-gene regulation, or potential horizontal pleiotropy. Nevertheless, CD33 remained the highest-priority protein candidate in the MR screen and was therefore taken forward for a conditional translational exploration. (Fig. 5e–g) Complete AlphaGenome, cross-analysis candidate-set, and six-variant functional annotation results are provided in Supplementary Tables S9a–S9h.

### 3.5 Structure-assisted prioritization of candidate CD33 small molecules

After genetic prioritization and regional functional interpretation, we conducted a post-prioritization translational exploration centered on CD33. This step was not intended to resolve the causal-gene assignment within the region; instead, because CD33 remained the highest-priority protein candidate at the MR stage, it translated that prioritization into an experimentally testable compound set. Known-drug and target-development sources were curated separately: contextual ChEMBL/DrugBank records did not automatically enter formal compound ranking, and antibodies and antibody–drug conjugates were excluded from docking. Formal experimentally supported small-molecule candidates came from the registered BindingDB/patent source set [44–45] and were ranked under the principle that experimental binding evidence took precedence over structural prediction.

The formal source set contained 322 standardized compound entities, of which 284 corresponded exactly to the GNINA production library and entered structure-assisted screening; the remaining 38 did not enter production. GNINA redocking across three cognate CD33 systems showed mixed pose-recovery performance, and production screening was fixed to the 5J06 chain-B receptor and 3′-sialyllactose pocket. The 284 production candidates were then screened with GNINA (Fig. 6a). Because redocking assessed native-pose recovery only and performance varied across reference systems, GNINA was consistently restricted to secondary ranking layered on existing experimental binding evidence.

**Figure 6.**
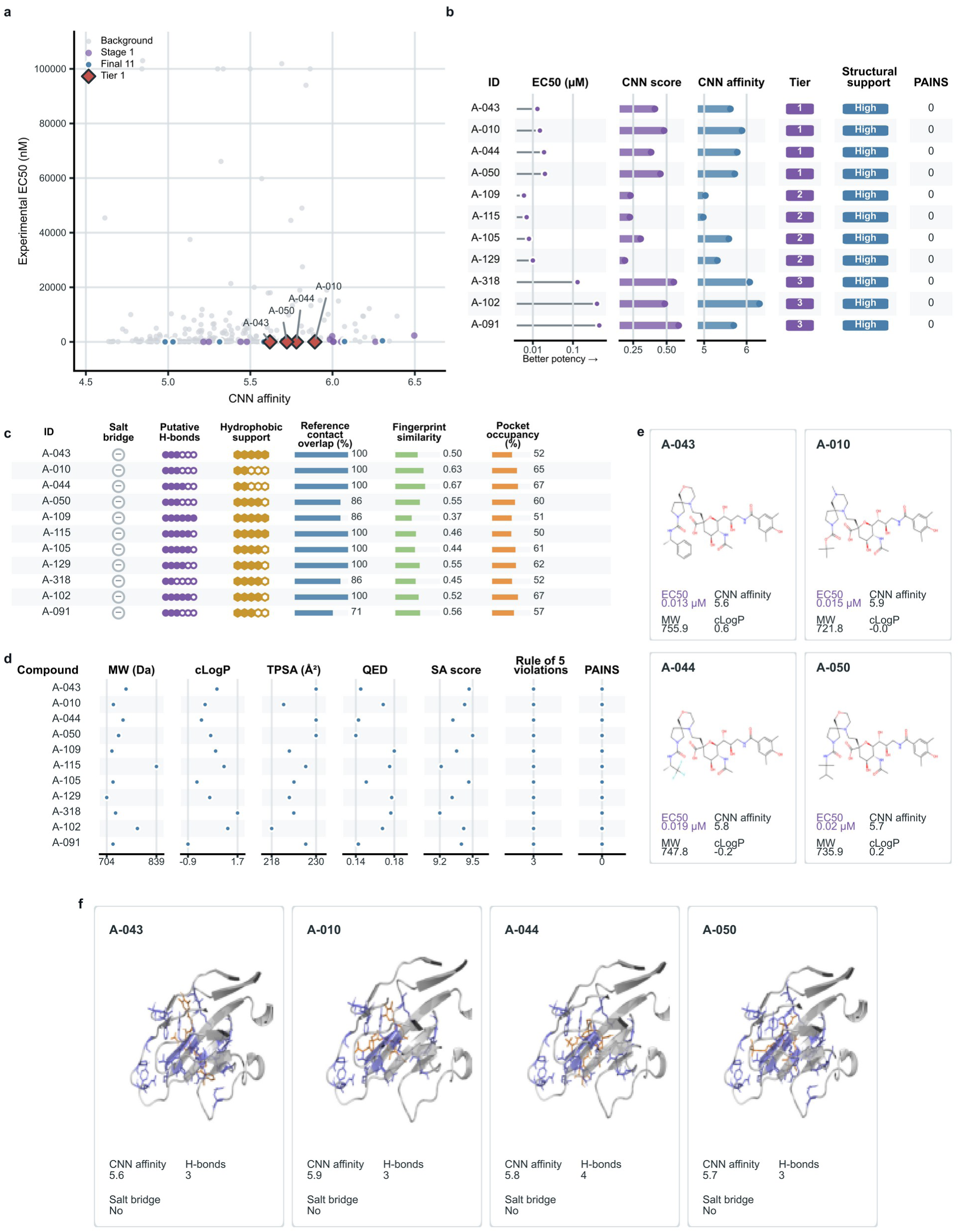
Experimental-binding-evidence-informed and structure-assisted prioritization of CD33 compounds. a, Reported recombinant human CD33 competitive-binding EC50 versus rank-1 GNINA CNNaffinity for the 284 production candidates. Background compounds, the 28 Stage 1 candidates, the final 11 retained candidates, and the four Tier 1 compounds are distinguished. b, Experimental potency, rank-1 CNNscore, rank-1 CNNaffinity, assigned tier, structural-support category, and PAINS status for the final 11 candidates. c, Structure-based descriptive metrics for the final 11, including salt-bridge status, putative hydrogen bonds, hydrophobic support, reference-contact overlap, fingerprint similarity, and pocket occupancy. d, Selected physicochemical and developability metrics for the final 11, including molecular weight, cLogP, TPSA, QED, synthetic-accessibility score, Rule-of-5 violations, and PAINS status. e, Chemical structures and selected experimental/computational properties of the four Tier 1 compounds A-043, A-010, A-044, and A-050. f, Predicted rank-1 GNINA poses for the same four Tier 1 compounds in the 5J06 chain-B/3′-sialyllactose production pocket, with CNNaffinity and salt-bridge detection status displayed. GNINA scores, contacts, and poses provide computational support only and do not represent measured affinity or experimentally confirmed binding modes. Reported competitive-binding EC50 likewise does not establish agonist/antagonist activity or pharmacological direction concordant with the MR effect. The circled “−” in panel c and “No” in panel f both indicate that no salt bridge was detected. PAINS, pan-assay interference compounds; QED, quantitative estimate of drug-likeness; TPSA, topological polar surface area.

Stage 1 integrated reported experimental binding potency, rank-1 GNINA performance, and valid pocket entry to retain 28 candidates from the 284 production compounds. Stage 2 applied prespecified structural-support, experimental-potency, and PAINS rules to assign Tiers 1–4, retaining 11 compounds in Tiers 1–3 and excluding Tier 4. Tier 1 comprised A-043, A-010, A-044, and A-050, with reported recombinant human CD33 competitive-binding EC50 values of 13–20 nM and relatively consistent computational structural support. Experimental measurements were taken from public patent/BindingDB records [44–45] and were not re-measured in this study. (Fig. 6b–f)

The final 11 compounds constitute a prioritized candidate set for subsequent experimental validation, with Tier 1 candidates showing the most consistent combined profile of reported binding potency and computational structural support. Complete redocking and compound-prioritization results are provided in Supplementary Tables S10a–S10e.

## 4 Discussion

We systematically screened predefined Ig-like plasma proteins against PTSD and related behavioral and neuropsychiatric traits. After the four MR associations meeting the prespecified screening criteria underwent the same post-screening robustness assessment, CD33–PTSD showed the most consistent support across instrument definitions and robust estimators and was advanced to regional genetic, brain molecular QTL, and functional analyses. Downstream findings did not establish a simple genetic-variant–CD33–PTSD mediation chain; instead, they localized the problem to the genetically complex CD33–SIGLEC region and yielded a limited set of prioritized candidate variants and experimental compound candidates. CD33 should therefore be regarded as the highest-priority PTSD-related plasma protein candidate among the predefined candidates in this study, rather than as a validated mechanistic target. Cross-method findings for CD33–PTSD and their interpretation boundaries are summarized in Supplementary Table S2.

This prioritization is biologically compatible with the neuroimmune context of PTSD. Large-scale genetic and molecular studies of PTSD implicate immune regulation, peripheral inflammation, and central neuroimmune processes in its complex biology. CD33, a myeloid cell-surface CD33-related Siglec, has been linked to monocyte and microglial function in other neurological diseases. Our scDRS/gsMap analyses likewise showed broad immune, vascular/stromal, neural, and barrier-related genetic contexts, but these are trait-level background findings and cannot identify the specific cell type through which CD33 might act in PTSD. At present, CD33-related myeloid immune regulation is better considered one experimentally testable mechanistic hypothesis. [1–2,4–6,57]

Regional genetic analyses further constrained interpretation of CD33–PTSD. Standard colocalization did not support a simple shared single variant; multi-signal SuSiE remained scientifically unresolved under the available finite external LD reference and summary–LD matching conditions; and SMR/HEIDI indicated regional heterogeneity from a different statistical perspective. Meanwhile, plasma CD33 pQTL shared strong regional signals with cortical CD33 eQTL and multiple sQTL molecular events, whereas these brain xQTLs did not show comparably robust sharing with PTSD. Together with AlphaGenome localization and functional annotation of the six candidate variants, involving neighboring genes such as SIGLEC9, KLK-family members, POLD1, and HAS1, these data support a framework compatible with complex LD, multiple signals, and multi-gene regulation.

At the translational level, the genetically prioritized CD33 candidate was converted into a directly testable compound set. Unlike a purely virtual screen, compound ranking was anchored first by previously reported recombinant human CD33 competitive-binding EC50 values [44], with GNINA providing only secondary structural support [46]; 11 candidates were ultimately retained. This analysis does not show that these molecules can modify PTSD and does not retrospectively strengthen the genetic evidence for CD33–PTSD. Competitive-binding EC50 does not define agonist/antagonist direction, consequences for myeloid-cell function, or concordance with the MR effect direction; orthogonal binding, receptor selectivity, and cellular functional assays are therefore required.

Several limitations remain. CD33–PTSD lacks independent genetic replication, and the relevant genetic datasets and LD reference are predominantly of European ancestry. Although multi-signal SuSiE was performed, it remained scientifically unresolved under the finite external LD reference; PGC SMR used a signed-Z/FREQ/NEFF working scale rather than original log-odds. Sharing between plasma pQTL and brain eQTL/sQTL does not itself establish brain CD33 protein regulation or disease mediation. AlphaGenome and some remote annotation resources have version/interface-level reproducibility limitations. In the structure-assisted screen, 284 of 322 standardized compounds entered production docking, while the remaining 38 were excluded because ETKDGv3 conformer generation failed during ligand preparation; static docking was additionally limited by inconsistent redocking performance across reference systems, and experimental binding potency and computational poses require further pharmacological and functional validation. Future studies should include independent and cross-ancestry validation, higher-resolution fine-mapping, brain/cerebrospinal fluid (CSF) or cell-type-specific protein QTL data, and experimental assessment of compound selectivity, cellular potency, and in vivo activity. Overall, this study proposes an experimentally testable CD33–PTSD hypothesis rather than a validated therapeutic target.

## Data availability

The datasets analyzed in this study were obtained from publicly available resources as described in the Methods and Supplementary Information. Processed data supporting the reported results, including source data underlying the figures and supplementary tables, are available to editors and reviewers during peer review upon request via a private repository and will be made publicly available in a permanent repository upon publication. Third-party datasets are not redistributed where redistribution is restricted and remain subject to the access conditions and terms of their original data providers.

## Code availability

Custom analysis and visualization code, together with associated reproducibility materials, are available to editors and reviewers during peer review upon request via a private repository. The code and reproducibility materials will be made publicly available in a permanent repository upon publication. Access to third-party software, databases, and restricted data resources remains subject to their respective licenses and terms of use.

## Ethics statement

This study was a secondary analysis of publicly available or previously published summary-level genetic, molecular-QTL, structural, and compound data. No new participants were recruited and no new human or animal samples were collected. Ethics approval and informed consent for the original human studies were obtained by the respective source studies under their original protocols. Because this study used no identifiable individual-level data, no new participant recruitment or intervention was undertaken.

## Acknowledgements

We thank the participants, investigators, consortia, and data providers of the original studies and public resources used in this work for generating and sharing the relevant genetic, molecular, and functional datasets. We thank Manting Fan for advice on aspects of the molecular docking analysis. The data used for the relevant analyses described in this manuscript were obtained from the GTEx Portal on August 5, 2026.

## Author contributions

Henghui Liu contributed to the conceptualization of the study and led the detailed development and implementation of the methodology, analytical code implementation, investigation, data curation, formal analysis, validation, visualization, and writing of the original draft and subsequent revisions. He also contributed to resource preparation and project administration. Shuyan Xie provided guidance on the implementation of selected analytical methods and contributed part of the experimental data and research resources. Zitao Chen contributed to conceptualization and methodology; provided guidance on the implementation of analytical methods, validation, formal analysis, and interpretation of results; and supplied part of the initial experimental data and research resources. He also contributed to project administration, supervision, manuscript review and editing, and provided suggestions on the interpretation of the findings and subsequent research directions. Qishan Wang contributed to conceptualization and methodology; provided guidance on the overall research direction, additional analytical approaches, validation and interpretation of results, and subsequent research directions; led supervision and overall project administration; and contributed to manuscript review and editing.

