## Supplementary Information for "Genetic and multi-omic prioritization implicates the CD33–SIGLEC locus in post-traumatic stress disorder"

### Contents

Supplementary Methods S1. Detailed scDRS and gsMap parameters

Supplementary Methods S2. Detailed parameters for bidirectional MR screening, internal evidence tiers, and robustness assessment

Supplementary Methods S3. Detailed parameters for CD33–PTSD regional genetic analyses

Supplementary Methods S4. Detailed parameters for BrainMeta cortical molecular-QTL sharing analyses

Supplementary Methods S5. Detailed parameters for AlphaGenome, regional localization, cross-analysis candidate sets, and functional annotation

Supplementary Methods S6. Detailed parameters for CD33 compound sources, redocking, and two-stage ranking

Reference numbering note

Supplementary Tables and support structure

### Supplementary Methods

#### Supplementary Methods S1. Detailed scDRS and gsMap parameters

scDRS: GWAS SNP P-value files for the 16 traits were processed at the gene level using MAGMA v1.10 custom [21] to generate gene ZSTAT values and assembled into a 16-trait gene-score input. scDRS 1.0.2 [22] compute_score used 1,000 control gene sets and filtered raw counts. Cell-type support required association_mcp≤0.05, assoc_mcz>0, and n_cell≥100 and was interpreted only as cell-type enrichment/prioritization rather than cell-type-specific causality. gsMap: under Python 3.12, gsMap 1.73.7 [23] quick mode was applied separately to two MOSTA E16.5 [24] mouse-embryo spatial sections with mouse–human ortholog mapping. Cross-section P values for the same trait–tissue pair were combined using the Cauchy method [25]. BH correction [26] was applied jointly across all retained trait–tissue Cauchy tests; FDR<0.05 was the primary threshold and FDR<0.10 was considered suggestive. Formal analysis records document the major versions of MAGMA, scDRS, and gsMap; some random seeds and complete environment locking were not recorded, while large atlases/resources are retained as registered external dependencies. [58–62]

#### Supplementary Methods S2. Detailed parameters for bidirectional MR screening, internal evidence tiers, and robustness assessment

Formal bidirectional MR analyses were based on unified results for 30 candidate proteins, with the same instrument-selection thresholds, LD-clumping settings, harmonization rules, estimator definitions, and diagnostic fields applied across proteins. Forward protein→trait analyses comprised 30×16=480 prespecified combinations, of which 476 were estimable, using pQTL instruments at $P<1\times10^{-5}$. Reverse trait→protein analyses covered 14 traits and yielded 420 estimable combinations, using GWAS instruments at $P<5\times10^{-8}$. PLINK2 [63] clumping used a 5,000-kb window, $r^2=0.01$, and p1=1; harmonization used TwoSampleMR [64] harmonise_data(action=2). Single-SNP analyses used the Wald ratio; multi-SNP analyses used IVW as the primary estimator [27] and additionally calculated MR-Egger [28], weighted median [29], simple mode, and weighted mode [30]. BH correction [26] was applied to the unified formal MR results within each MR direction×method family. Formal MR output defined three supporting evidence categories: High, Moderate, and Suggestive; remaining results were labeled Not supported, and records to which the classification did not apply were labeled N/A. High required IVW BH-FDR<0.05, MR-Egger intercept P≥0.05 or unavailable, and IVW Cochran-Q P≥0.05 or unavailable. Moderate required IVW P<0.05 and MR-Egger intercept P≥0.05 or unavailable, together with either IVW BH-FDR<0.10 or at least two formal MR methods with P<0.05 and concordant effect direction. Suggestive included results with IVW P<0.05 that did not meet all High or Moderate criteria. Results satisfying the internal High definition are reported consistently in the main manuscript and the core CD33–PTSD evidence matrix as "associations meeting the prespecified screening criteria"; this reporting label does not alter the internal classification, screening thresholds, BH-correction family, or result assignment. The four pairs meeting the prespecified screening criteria then entered post-screening robustness/sensitivity analyses: the primary threshold $P<1\times10^{-5}$, the stringent threshold $P<5\times10^{-8}$, and cis defined as the GRCh37 target gene±1 Mb. Each new instrument set was reclumped and reharmonized under the same rules. Analyses with 0 SNPs were non-estimable, those with 1 SNP used the Wald ratio, and multi-SNP sets used IVW and applicable supplementary estimators. Heterogeneity, Egger-intercept, and primary leave-one-out results were reused/verified, with additional cis leave-one-out analysis. MR-PRESSO [31] required ≥4 SNPs, 1,000 simulations, and alpha=0.05; MR-RAPS [32] was attempted for the primary sets and eligible cis sets. Steiger filtering was not performed where reliable variance metadata were unavailable. This component was a robustness/sensitivity assessment rather than independent replication. Only BIP could be expressed on an OR scale; PTSD and BeerPref remained on the beta scale, while the available pQTL headers did not permit verification of one-SD scaling. Advancement from the four retained pairs was not governed by a second formal significance threshold. Instead, all four pairs were compared within the same robustness framework, and CD33–PTSD was advanced because it showed the most consistent support across alternative instrument definitions and applicable robust estimators; this was a qualitative post-screening prioritization rather than independent replication or a prespecified binary gate. The current fixed-RNG reconstruction environment was standardized and records R 4.5.2, MASTER_SEED=20260905, RNGkind, and the pair-level seed map, with exact reproducibility verified by clean-session RUN A/RUN B; only some historical R/package patch information and the exact binary/version of the upstream external PLINK2 were not fully recovered.

**Supplementary Methods S3. Detailed parameters for CD33–PTSD regional genetic analyses**

The formal standard single-signal colocalization analysis was restricted to plasma CD33 pQTL–PGC PTSD and used coloc.abf [33] within GRCh37 regions of ±250 kb, primary ±500 kb, and ±1 Mb, with at least 50 shared SNPs per window. $p1=p2=1\times10^{-4}$; the primary p12 was $1\times10^{-5}$, with $1\times10^{-6}$, $5\times10^{-6}$, $1\times10^{-5}$, and $5\times10^{-5}$ evaluated. The QC grid covered MAF thresholds of 0/0.01, INFO thresholds of 0/0.8, indel/palindrome handling, and an ambiguity cutoff of 0.42 for palindrome MAF. H4≥0.8 was classified as strong sharing, H4≥0.5 as moderate sharing, H3≥0.8 as distinct signals, and H4/H3≥3 was recorded as relative dominance; these are posterior interpretation rules rather than frequentist P values. The PGC outcome working scale was constructed from signed Z/FREQ/NEFF and cannot be interpreted as original log-odds/OR or exponentiated as original log-odds/OR; standard colocalization is not fine-mapping. Multi-signal SuSiE-RSS [34–35] analyses used 1000 Genomes Phase 3 EUR external LD [65] (B=489), MAF≥0.01, a frequency-mismatch cutoff of 0.10, and required ≥100 matched SNPs per window. To enable direct comparison across models, four models were compared using the same matched SNP set and identical LD ordering: Model A (ordinary/reference baseline), Model B (finite-only, R_finite=489), Model C (primary mismatch-aware, R_mismatch=eb), and Model D (secondary eb_mix sensitivity). All four models used L=10, max_iter=50, coverage=0.95, min|r|=0.50, check_prior=TRUE, residual_variance=FALSE, and refine=FALSE. The reliability gate included flags for |Z| >40, frequency mismatch 0.10, summary–LD s warning/severe thresholds of 0.10/0.50, kriging logLR≥2 with |

Z|≥2, suspicious proportion 0.001, r/B≤0.20, prior variance>100, PIP≥0.999, and ≥2 singleton pathologies. coloc.susie [36] was interpreted only when the primary Model C for both datasets within the same window was RELIABLE. PIP/CS/PP.H4 outputs that did not pass this gate were retained as diagnostic records and were not used as positive or negative evidence of sharing. target-SMR/HEIDI [37] and SMR-multi [38] used the same PGC working scale and EUR LD B=489: CD33±1 Mb, pQTL $P<1\times10^{-5}$, F>10, target independence $r^2<0.01$, allele-frequency difference≤0.20, and ambiguous-palindrome filtering at EAF 0.42–0.58; peqtl_smr=$1\times10^{-5}$, peqtl_heidi=$1.57\times10^{-3}$, HEIDI LD 0.05–0.90, min 3/max 20, method 1. target-SMR used BH correction across the tested targets, nominal alpha=0.05, and HEIDI P<0.01 as a warning against a simple shared model; failure to reject by HEIDI does not prove colocalization. The prespecified top cis-pQTL was rs2455069/19_51728641. SMR-multi was a single set-level test with internal multi-SNP LD=0.10 and does not constitute SNP-level significance, replication, or mediation evidence. SMR was executed using a locally compiled binary; the existing retained records do not uniquely recover the exact release/commit that was run.

**Supplementary Methods S4. Detailed parameters for BrainMeta cortical molecular-QTL sharing analyses**

BrainMeta v2 [39] cortical analyses comprised two non-combined families of pairwise colocalization. The first family tested plasma CD33 pQTL against one cortical CD33 eQTL context and nine sQTL molecular events; the second family separately tested the corresponding cortical eQTL/sQTL events against PGC PTSD. The region was defined as GRCh37 CD33 gene endpoints±1 Mb (chr19:50,728,320–52,747,115). Each comparison independently used coloc.abf [33] with $p1=p2=1\times10^{-4}$, primary $p12=1\times10^{-5}$, $1\times10^{-6}/1\times10^{-4}$ sensitivity analyses, MAF≥0.01, and at least 100 matched SNPs. The nine sQTL records are molecular events rather than nine independent genetic signals; the two families do not constitute a three-variable model, mediation analysis, tissue-directionality inference, or fine-mapping. The complete 32-GB BrainMeta source archive is retained as an external dependency, while the analysis repository retains directly analyzable derived data and metadata.

**Supplementary Methods S5. Detailed parameters for AlphaGenome, regional localization, cross-analysis candidate sets, and functional annotation**

AlphaGenome [40] used the local Python SDK v0.6.1 with a 131,072-bp sequence context. Variant REF/ALT alleles were checked in GRCh37/hg19 using Ensembl REST as the primary source; UCSC hg19 was used as fallback only when the Ensembl query failed, was unresolved, or showed a REF/ALT mismatch. Raw scores were ALT−REF and were direction-flipped when the pQTL effect allele was REF. High priority required resolved alignment, direction concordant with the pQTL, and a within-group percentile rank≤0.05 for |effect-allele score|; after excluding high-priority variants, moderate priority required percentile≤0.15. This rule was used only for functional prioritization. The exact server-side AlphaGenome model/API backend was not recorded in the available analysis materials. Nearest gene/TSS, target-gene body, strand-aware promoter, CD33±500-kb proximity, and regulatory-region overlap were then recorded in GRCh37. The promoter was defined as 2,000 bp upstream/200 bp downstream of the TSS: [TSS−2000, TSS+200] on the positive strand and [TSS−200, TSS+2000] on the negative strand, inclusive of endpoints. The CD33–SIGLEC locus was predefined as chr19:51,000,000–52,500,000; gene proximity was not treated as target validation. The strict four-set membership intersection comprised formal CD33–PTSD MR instruments, all members tested in target-SMR, verified effective SMR-multi members, and CD33 AlphaGenome high-priority members. target-SMR membership did not require SNP-level significance, and SMR-multi membership likewise did not denote SNP-level significance. The six fixed candidates were rs12973028, rs3212815, rs117268623, rs10426620, rs2075803, and rs181390202. This intersection represents membership convergence only and is not evidence of colocalization, mediation, a credible set, or shared causality. VEP [41] consequences and source-defined significant single-tissue GTEx v8 [42] eQTLs were further used to annotate the six SNPs; all six fixed candidates completed GRCh37→GRCh38 identity mapping and REF/ALT verification for downstream VEP/GTEx integration, with no additional project-level significance threshold introduced. Non-return by a provider endpoint was not interpreted as absence of a regulatory effect. Saved formal RegulomeDB v2 [43] exports were used for all six rsIDs, retaining source rank/probability/evidence without rescoring; detailed stable-version metadata are incomplete. RegulomeDB was used only as descriptive regulatory annotation and not as independent functional validation or evidence establishing causality. Targeted six-SNP×CD33 GTEx queries were also performed: eQTLs were queried through nominal dyneqtl across 49 v8 tissues, with source-significant exact-match rows flagged separately; sQTLs were queried only through exact variant–CD33 significance endpoints, without exhaustive enumeration of nominal sQTLs in the absence of a prespecified splicing phenotype. NES retained the GTEx ALT direction and was not flipped to the MR effect-allele direction.

**Supplementary Methods S6. Detailed parameters for CD33 compound sources, redocking, and two-stage ranking**

The formal CD33 small-molecule/carbohydrate source set and the docking-ready set were registered separately. Conservative entity standardization used names, canonical isomeric SMILES, InChIKey, and quantitative activity. The formal 322 unique InChIKey entities came from the registered BindingDB/patent source collection [44–45]; ChEMBL [47] and available licensed DrugBank records [48] were searched separately for known CD33-targeting drugs/compounds, but no CD33-targeting small-molecule drug meeting the study inclusion criteria was identified. These search records were not added to the 322 entities or to docking ranking, and antibodies and antibody–drug conjugates were excluded from small-molecule docking. TTD [49], BindingDB, Thera-SAbDab [50], ClinicalTrials.gov, and related sources were used separately to describe the target-development landscape; they did not define the count of 322, the 284-member production set, or Stage tiering. Complete licensed DrugBank data were unavailable. Among the 322 standardized entities, 284 had exact entity-level one-to-one correspondence with the production candidates through standardized compound identifiers, while 38 did not enter production. All 38 were excluded at the ligand-preparation stage because ETKDGv3 [51] conformer generation failed. Before formal GNINA redocking validation [46], AutoDock Vina [66] was first used to redock the three reference systems 7AW6/FVP [52], 5J06/3′-sialyllactose [53], and 5J0B/6′-sialyllactose [54]; none met the prespecified pose-recovery requirement, so Vina was not used for subsequent production screening and GNINA was instead used to validate redocking in the same three systems. GNINA redocking then used 7AW6/FVP, the Neu5Ac–Gal core of 5J06 3′-sialyllactose, and the Neu5Ac–Gal core of 5J0B 6′-sialyllactose on CPU with GNINA v1.0.3, cnn_scoring=rescore, exhaustiveness=16, 20 modes, a 6-Å autobox, and seeds 20260809/20260909/20261009, respectively. RMSD was a heavy-atom, symmetry-aware RMSD in the receptor coordinate frame without ligand fitting: EXCELLENT=rank1<2 Å; otherwise ACCEPTABLE=best<2 Å; PARTIAL=best<3 Å; otherwise FAIL. The unmodeled reducing-end glucose in the 5J06/5J0B crystal ligands was excluded from RMSD. Redocking assessed pose recovery only. Formal production screening used the 5J06 chain-B receptor/3′-sialyllactose pocket on CPU with GNINA v1.0.3, box center=(−5.3307,10.5751,−0.2165) Å, size=(24.5570,18.0790,20.3900) Å, exhaustiveness=16, 10 modes, seed=20260909, and rescore.

Stage 1 assigned all 284 production candidates to Groups A/B/C/D in a fixed sequential order, stopping at the first satisfied condition. Experimental-activity thresholds used for grouping were Q2.5=14 nM, Q25=101 nM, and Q75=3,000 nM; structural-score thresholds were the median rank-1 CNNscore across all 284 candidates (0.308967859), the median rank-1 CNNaffinity (5.355744605), CNNscore Q90=0.468628269, and CNNaffinity Q75=5.628250005. Group A (concordant high priority) required an exact numeric CD33 EC50≤101 nM, rank-1 CNNscore≥0.308967859, rank-1 CNNaffinity≥5.355744605, a valid rank-1 pose, and entry of the rank-1 pose into the reference 3′-sialyllactose pocket. Group B (experimentally strong, dockingdiscordant) was evaluated only after failure to meet Group A and required an exact numeric CD33 EC50≤14 nM, without requiring CNNscore, CNNaffinity, pose validity, or pocket entry; therefore, candidates with EC50≤14 nM that also met the Group A structural conditions remained in Group A. Group C (structurally strong, moderate experimental activity) was evaluated only after failure to meet Groups A/B and required an exact numeric CD33 EC50 with 101<EC50≤3,000 nM, rank-1 CNNscore≥0.468628269, rank-1 CNNaffinity≥5.628250005, a valid rank-1 pose, and entry into the reference pocket. All remaining candidates were assigned to Group D (not selected at Stage 1). Stage 1 retained Group A∪Group B∪Group C, whereas Group D did not advance; the frozen result was A=15, B=7, C=6, D=256, for 28 retained candidates.

Pocket entry was defined as follows: the reference pocket comprised receptor heavy atoms in 5J06 lying within ≤5.0 Å of any heavy atom of the native 3′-sialyllactose; a candidate rank-1 pose was considered to enter the pocket if at least one ligand heavy atom lay within ≤4.5 Å of these pocket atoms. Groups A and C required both a valid pose and pocket entry, whereas Group B did not require these structural conditions. Best Vina and rank-1 Vina were retained only for auxiliary reporting and did not participate in A/B/C/D assignment; docking-protocol parameters conferred no selection advantage in Stage 1 grouping. Molecular properties, PAINS, pharmacology, selectivity, cellular activity, compound series, and development stage did not enter the Stage 1 rules.

High structural support required rank-1 occupancy≥0.50, reference overlap≥0.25, top-3 stability≥0.40, pocket fraction≥2/3, and an ARG119 contact; moderate support required occupancy≥0.25, overlap>0, stability≥0.25, and pocket fraction≥2/3. Stage 2 rules were: Tier 1=Group A+EC50≤20 nM+high/moderate support+PAINS=0 [55]; Tier 2=Group B+EC50≤10 nM; Tier 3=Group C+EC50≤500 nM+high/moderate support; all remaining candidates were Tier 4 and were not retained. No weighted final score was used; properties were background information only, and candidate-level selectivity/cellular activity was unavailable.

### Reference numbering note

Numerical reference citations in the Supplementary Methods follow the Reference list in the main manuscript. To avoid duplication, the Supplementary Information does not repeat the full references already provided in the main manuscript.

### Supplementary Tables and support structure

Complete results are assembled from the formal result files in Supplementary_Tables.xlsx. The Supplementary Methods describe how analyses were performed, whereas the Supplementary Tables provide the complete analytical results. Supplementary Table S2 summarizes cross-method evidence and interpretation boundaries for CD33–PTSD; its purpose is to present findings and permitted interpretations side by side by analytical domain, rather than treating MR, colocalization, SMR, brain xQTL, functional annotation, and docking as independent votes or calculating a cross-method aggregate score. The workbook contains 41 worksheets: S1, S2, S3, S4a, S4b, S5, S6a–S6h, S7a–S7l, S8a–S8b, S9a–S9h, and S10a–S10e.

| Supplementary Table | Module | Required content and reporting boundary |
|---|---|---|
| Table S1 | Research inputs and data sources | Phenotype definitions, sample sizes, effect/statistical scales, genome builds, ancestry, and source/access information for the 16 traits, pQTL, PGC PTSD, BrainMeta, MOSTA/LD, and |

| Supplementary Table | Module | Required content and reporting boundary |
|---|---|---|
| | | other formal resources. |
| Table S2 | Cross-method evidence and interpretation boundaries for CD33–PTSD | Summarizes the key findings and permitted interpretation boundaries for MR, disease colocalization, SMR/HEIDI, SMR-multi, brain xQTL, functional annotation, and translational exploration; no star rating, cross-method aggregate score, or majority vote is used. |
| Table S3 | Candidate protein definition | The 184 Repetto Olink assays yielded 178 unique and unambiguous gene symbols after unambiguous assay–gene mapping and gene-symbol deduplication; 30 candidate proteins were then fixed according to IPR003599 membership. |
| Tables S4a–S4b | Trait-context analyses | S4a: complete scDRS cell-type results; S4b: complete gsMap trait–tissue Cauchy/BH results. Both provide trait-level background only and do not participate in protein–trait candidate prioritization. |
| Table S5 | Bidirectional MR screening | Complete unified forward/reverse MR estimates for the 30 proteins, including method, nSNP, effect, SE/P, BH-FDR, and pair-level internal classification (High, Moderate, Suggestive, Not supported; separately marked when not applicable). The complete 480-cell forward screening grid, including four non-estimable combinations, is provided in Source Data Fig2E_MR_screen. In manuscript-facing reporting, High-class results are uniformly described as “associations meeting the prespecified |

| Supplementary Table | Module | Required content and reporting boundary |
| --- | --- | --- |
| | | screening criteria”. |
| Tables S6a–S6h | Post-screening MR robustness | S6a: primary MR reference–rerun reproducibility check; S6b: MR estimates under alternative instrument definitions including cis-only, stringent-all, and stringent-cis; S6c: integrated post-screening MR sensitivity summary; S6d: instrument composition; S6e: primary/cis-only leave-one-out; S6f: MR-PRESSO; S6g: MR-RAPS; S6h: heterogeneity/horizontal-pleiotropy diagnostics including Cochran-Q, MR-Egger intercept, and Steiger status. This module is a common post-screening robustness assessment rather than independent replication. |
| Tables S7a–S7l | CD33–PTSD regional genetic analyses | S7a: formal standard single-signal colocalization windows/priors/QC and PP0–PP4; S7b: SuSiE regional input summary; S7c: summary–LD diagnostics; S7d: finite-reference diagnostics; S7e: LD matrix QC; S7f: four-model SuSiE fit summary; S7g: reliability QC decisions; S7h: standard-coloc comparison calculated on the SuSiE-matched SNP set; S7i: PIP/credible-set diagnostics; S7j: coloc.susie signal-pair and p12-prior sensitivity; S7k: target-SMR/HEIDI; S7l: SMR-multi set-level results. SuSiE-related outputs in S7b–S7j are reported as diagnostic and scientifically unresolved under the current external-LD conditions; the matched-set standard-coloc values in S7h are not required to equal the formal standard-coloc |

| Supplementary Table | Module | Required content and reporting boundary |
| --- | --- | --- |
| | | values in S7a. |
| Tables S8a–S8b | Brain molecular-QTL sharing | S8a: pairwise colocalization of plasma CD33 pQTL with cortical CD33 eQTL/nine sQTL molecular events; S8b: pairwise colocalization of the corresponding cortical eQTL/sQTL with PTSD. Both families report event-level PP0–PP4 and p12 sensitivity and are interpreted as two independent analysis families rather than combined into a mediation chain. |
| Tables S9a–S9h | Variant prioritization and functional annotation | S9a: AlphaGenome long-format sequence-functional prediction and locus annotation; S9b: membership matrix for MR instruments, target-SMR, SMR-multi, and AlphaGenome high-priority sets and the six candidates in the strict four-set intersection; S9c: VEP, general GTEx-source annotation, and RegulomeDB annotation for the six candidates; S9d: eQTL summary for the targeted six-SNP×CD33 queries; S9e: targeted CD33 sQTL summary; S9f: targeted CD33 sQTL endpoint/query results; S9g: targeted GTEx query QC; S9h: tissue-level targeted CD33 nominal eQTL results. Non-return/failure states in targeted queries are not interpreted as biological negatives, and GTEx NES retains the source ALT direction. |
| Tables S10a–S10e | Translational compound analyses | S10a: GNINA redocking across three CD33 reference systems with rank-1/best RMSD and pose-recovery category; S10b: identity, status, and exclusion |

| Supplementary Table | Module | Required content and reporting boundary |
|---|---|---|
| | | reasons for the 322 source entities, 284 production members, and 38 entities not entering production; S10c: experimental binding information, GNINA results, and Stage 1 A/B/C/D grouping for the 284 production candidates; S10d: structural-support, contact, and pocket metrics for the 28 Stage 1 candidates; S10e: physicochemical/structural-alert information, Tier 1–4 assignment, and final retention of 11 candidates among the 28 Stage 1 candidates. |

Supplementary Figures: the main manuscript currently does not depend on any Supplementary Figure. Additional supplementary figures should not be created solely for formal completeness unless a main-text panel is moved during subsequent layout preparation.
